# Rapid slide-free histopathology via fluorescent layer-adaptive subcellular imaging: clinical-accessible platform for diverse tissues

**DOI:** 10.1101/2025.04.07.25325427

**Authors:** Xiaoguang Guo, Xingguang Chen, Fanhao Qiu, Yaolong Li, Yan Wu, Zhengxia Wang, Yingjun Zhang, Zhen-Li Huang

## Abstract

Intraoperative pathology remains constrained by ice crystal artifacts in frozen sections and the high cost of emerging slide-free optical methods. Here, we introduce FLASH-Path, a rapid slide-free technique enabling subcellular-resolution imaging of centimeter-scale tissues in 10 minutes. By replacing mechanical thin sectioning or optical thin-layer excitation with thin-layer (≤10 µm) fluorescent labeling using commercially available probes, FLASH-Path achieves artifact-free visualization of diverse tissues (e.g., fat, lymph nodes) incompatible with conventional frozen sections. The method integrates with retrofitted clinical fluorescence microscopes or manual observation, ensuring adaptability across resource settings. Fluorescence images are computationally transformed into H&E-like histopathology without ice crystal artifacts or hidden risks from generated images. In colorectal cancer validation, FLASH-Path outperformed frozen sections in speed and imaging area. FLASH-Path enhances clinical accessibility, image traceability, and cost-effectiveness, providing new opportunities for the clinical application and dissemination of slide-free pathology.

## Introduction

Global cancer burden reached 20 million new cases and 9.7 million deaths annually in 2022, projected to surge to 35 million new cases by 2050 ^1^. Surgical resection remains the cornerstone of treatment, where complete tumor removal with maximal preservation of normal tissue critically determines patient survival and quality of life ^2^. Intraoperative pathology plays a pivotal role by examining tissue sections or images to detect structural and cellular atypia ^3^, guiding surgery and tumor margin identification. For the example of colorectal cancer (CRC), the second most common cancer in China ^4^, precise localization of surgical margins— particularly in mid-low rectal cancer—is essential for prognosis and preserving anal sphincter function ^5^. However, assessing the extensive distal and circumferential margins, which contain significant fat, poses challenges for current intraoperative frozen section pathology and emerging slide-free pathology methods. This often hinders fast, accurate, and comprehensive evaluation within the limited surgical time, increasing the risk of secondary surgery and short-term recurrence for many patients ^6^.

Current frozen section pathology, while important for intraoperative diagnosis, faces several limitations ^7^: (1) Technical complexity requiring skilled technicians and high-cost equipment maintenance; (2) Sample constraints (≤15×15 mm per section, 20-minute processing) that exclude lymphocytic tumors, calcified tissues, and adipose-rich specimens ^8^ due to low freezing points-induced ice crystal artifacts ^9^; (3) Morphological distortion from ice crystals and thick-section detail loss ^10^; (4) Sample non-reusability necessitating repeated sampling. These limitations force large surgical margins (e.g., CRC) into fragmented analysis, increasing diagnostic delays and risk of error.

In recent years, advances in imaging technology have revolutionized traditional histopathology, with innovative optical imaging methods such as ultraviolet surface-excitation microscopy (MUSE) ^11^ and ultraviolet photoacoustic microscopy (UV-PAM) ^12^ enabling high-contrast, slide-free pathology ^13^. MUSE uses ∼280 nm UV light for real-time 3D contour scanning to image, while UV-PAM uses a 266 nm pulsed laser to image untreated bone samples.

Both methods use unsupervised deep learning to convert grayscale images into images that resemble stained Hematoxylin & Eosin (H&E) histopathology, as is customary for pathologists. However, the slide-free imaging technology based on deep ultraviolet light source often depends on the absorption characteristics ^14^ or autofluorescence characteristics ^15^ of the sample, and the spatial heterogeneity of these optical characteristics may affect the recognition of some structural details. Consequently, some pathologists remain skeptical of virtual staining due to its poor interpretability, which raises concerns about hidden risks from generated images ^16^. In addition, the high cost of deep UV microscopes and detectors, as well as the long period of specialized training, hinders their adoption in hospitals that are sensitive to medical risk and price ^17^. For intraoperative rapid pathology to succeed, clinical accessibility hinges foremost on diagnostic accuracy (minimizing medical errors) ^18^ and secondarily on a balanced integration of speed, interpretability, comprehensiveness, user-friendliness, affordability and artificial intelligence ^19^. While promising, most emerging slide-free pathology technologies struggle to concurrently meet these rigorous criteria.

In-depth analysis reveals that the rapid acquisition of structural and cellular heterogeneity information of lesion tissues is a core requirement for intraoperative pathological diagnosis ^20^. Based on this, we propose a novel technique: Fluorescent Layer-adaptive Artifact-free Subcellular Histopathology via Pixel-preserving H&E Image Conversion (FLASH-Path). This technique uses the new concept of thin-layer staining, replacing the popular idea of thin-layer physical sectioning (used in traditional H&E histopathology) and thin-layer optical excitation (used in many recent slide-free pathology techniques). Firstly, commercial fluorescent probes are used to stain cell nucleus (DAPI) and membrane (DiD) with a thickness of ∼10 μm (equivalent to the thickness of a frozen section sample ^7^); Secondly, image scanning is performed using a fluorescence microscope equipped with automatic XY stage (area scanning) and motorized focus for extended depth of field (EDF) function ^21^, which can be easily upgraded from clinical fluorescence microscope; At last, through the mapping calculation of the color channels, the original fluorescence images (hereafter called DD images, since they are from DAPI and DiD staining) can be linearly transformed into the customary H&E staining pattern (i.e., the DD-HE image), without loss of resolution or hidden risks from generated images. The DD-HE image not only obtains information on tissue structure and cell morphology, but also obtains information on subcellular structures such as cell membranes and nuclei, facilitating subsequent cell segmentation and identification ^22^.

We validated FLASH-Path using both mouse colon and human CRC samples, demonstrating its clinical accessibility, image traceability, and cost-effectiveness. Notably, FLASH-Path delivers H&E-like image quality, surpassing frozen sections by enabling superior visualization of both fat and lymphocytes without the inherent drawbacks of frozen section pathology ^23^. This is accomplished while maintaining rapid processing of 15×15 mm samples within 10 minutes (including sample preparation, optical imaging and image transformation), and breaks through the sample size limitations of traditional sectioning molds to enable fast and complete imaging of large samples of at least 35×45 mm, and the size is only limited to the travel range of the XY stage. Image traceability is ensured through a color channel mapping algorithm that integrates fluorescent and white light image data with associated biological information, establishing a physically grounded and reproducible workflow. Furthermore, the FLASH-Path system allows for both on-site manual adjustment and observation using a clinical fluorescence microscope, as well as large-area scanning imaging through easy system upgrades.

This dual capability not only enhances affordability for primary healthcare facilities, but also provides a strong foundation for digital image archiving, remote consultation and artificial intelligence (AI) applications in histopathology ^24^. Importantly, the technology preserves tissue cell structures and bio-antigenic components during tissue processing and image acquisition, allowing for valuable downstream analyses such as immunohistochemistry, immunofluorescence, and gene detection, highlighting its high practicality, compatibility, and significant potential for widespread implementation.

## Results

### 1. FLASH-Path Principle

The optical system of FLASH-Path integrates two lasers and a motorized XYZ sample stage, supplemented by the optical components of a conventional fluorescence microscope and an EDF scanning program (see Fig. 1a for the optical path design; see Methods 4 for details of the image acquisition method). The key to sample processing lies in the selection of fluorescent dyes and the precise control of staining thickness (see Fig. 1b for penetration curves; see supplementary Fig. S3 for results of confocal microscope images). The construction of the system and the thin-layer staining laid the foundation for the subsequent EDF scanning (see Fig. 1c for the EDF process; see supplementary Fig. S2 for the image stitching technique).

**Fig. 1.**
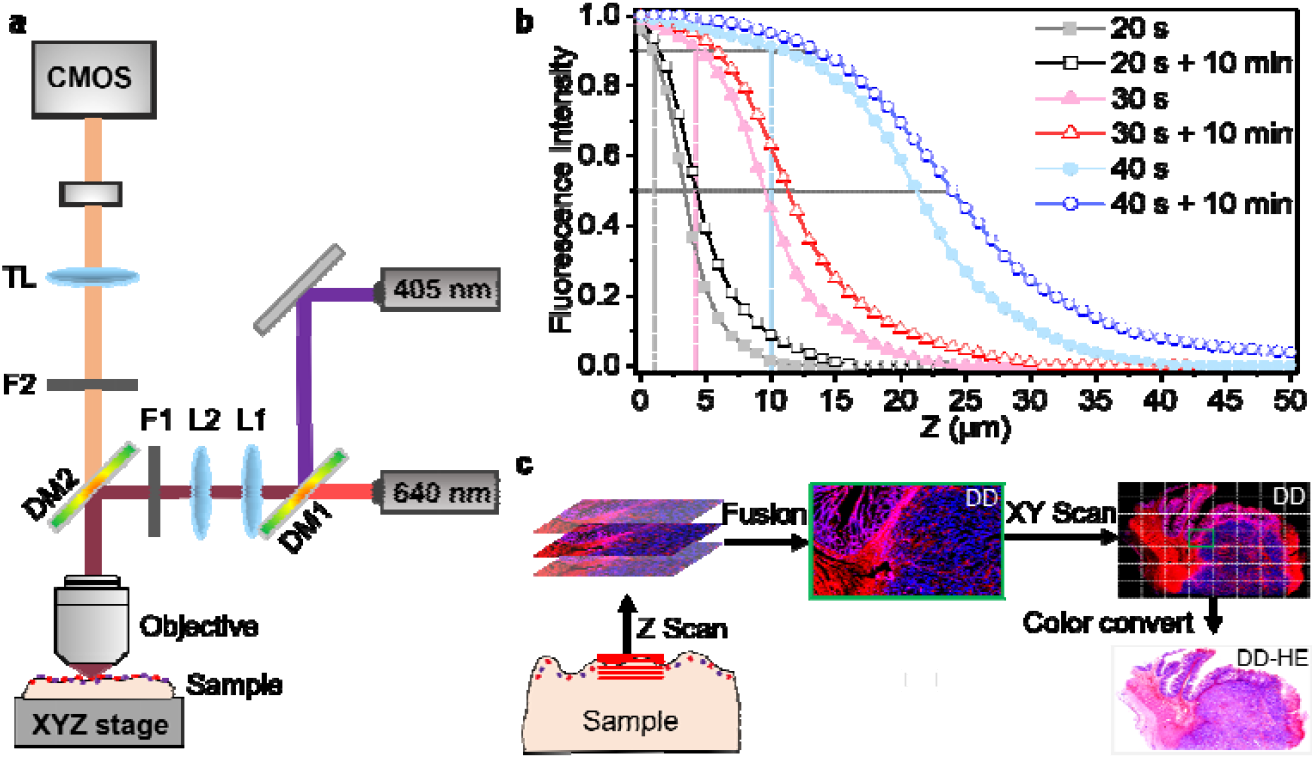
Principle of FLASH-Path. **a**. The optical system. The two excitation lasers are spatially combined using a dichroic mirror (DM1), modulated in spot size by a lens group (L1 and L2), purified with an excitation filter (F1), and reflected to the Objective with a dichroic mirror (DM2). The collected fluorescence by the Objective passes the dichroic mirror (DM2), purified with an emission filter (F2), and finally focused by a tube lens (TL) onto the color camera (CMOS). For detailed information of the optical system, see Image acquisition in the Methods section. **b**. The dependence of normalized fluorescence intensity on the sample thickness and stained duration (20, 30 and 40 s). The same samples were kept undisturbed for 10 minutes prior to undergoing a second round of optical imaging. Scanning parameters: Z-axis step size = 1 μm, a total of 51 images were collected. **c**. The schematic diagram of the imaging process (See Supplementary Fig. S2 for a more detailed description).

In the selection of fluorescent dyes, we took the tumor cell heterogeneity identification features (e.g., nuclear morphology and nuclear-to-cytoplasm ratio) as the starting point, and paid special attention to the specific staining of cell nuclei and cell membranes. After a series of pre-experimental comparisons, we found that DAPI showed the best specificity and contrast for nuclear staining and DiD for cell membrane staining. Although rhodamine ^25^, acridine orange ^26^ and other membrane staining dyes were available, they tended to stain both the cytoplasm and intercellular matrix, resulting in blurred cell boundaries, which was not conducive to the determination of the nucleus-to-plasma ratio (a strong predictor of malignancy) and subsequent cell segmentation.

The control of the staining thickness is the key to the technique: DAPI can be excited by a short wavelength light source, which means the illumination will be limited in a thin layer; whereas DiD needs to be excited by a 640 nm red laser, which can penetrate the tissues several millimeters deep ^27^, and therefore the staining depth of DiD must be precisely controlled. Fig. 1b shows the relationship among normalized fluorescence intensity, staining duration, and the staining thickness of the sample, and supplementary Fig. S3 shows more experimental details. By controlling the residence time of the staining solution on the sample surface, we were able to regulate the penetration depth of the probe molecules, and the optimal staining thickness was expected to be 4-10 μm (the thickness threshold for physical sectioning), which was achieved in 20-30 s of staining (see Methods for details of the procedure).

The imaging system uses a high-resolution color camera with a pixel size of 2.4 μm and a peak quantum efficiency of 84% at 535 nm, which enables efficient acquisition and differentiation of dual-channel signals while ensuring high-quality tissue imaging. For surface imaging of slide-free samples, we introduced a motorized XYZ stage in the imaging system (Fig.1a). This stage enables micrometer-level 3D scanning near the sample surface ^28^. Additionally, it matches the system’s depth of field. By combining this XYZ stage with multifocal image fusion, we achieve panoramic fluorescence imaging of tissue samples with no upper limit on the sample size. Detailed information on system accessories can be found in the Image Acquisition of the Methods section. The sample scanning and image processing flow is shown in Fig. 1c.

### 2. Quantitative Equivalence Between FLASH-Path (Slide-Free) and FFPE-HE (Sectioned) Imaging Platforms

The quality of histologic images is crucial in diagnostic pathology, since it affects the accuracy of disease diagnosis by pathologists ^29^. Among them, the sharpness, contrast, color fidelity and noise control of histological structures (epithelial, vascular, lymphatic, muscle and cancerous tissues, etc.) are the basic morphological requirements for all disease diagnoses.

For evaluating the image quality of FLASH-Path presented by DD-HE images, in this paper, human CRC samples were firstly subjected to DD staining and slide-free imaging to obtain fluorescence images (Slide-free DD, Fig. 2a), which were converted to DD-HE images (Fig. 2b). The tissues used in FLASH-Path were processed with standard formalin-fixed paraffin-embedded sectioning (FFPE) and imaged using bright-field microscopy, and thus presented conventional FFPE-HE (Hematoxylin & Eosin histopathology with formalin-fixed paraffin-embedded sectioning) images for comparison (Fig. 2c). It should be noted that the FFPE-HE and DD-HE images shown in Fig. 2 are histologic images at different tissue depths but in close proximity to each other. The protocols for sample processing, imaging and color conversion are presented in the Materials and Methods section accordingly.

**Fig. 2.**
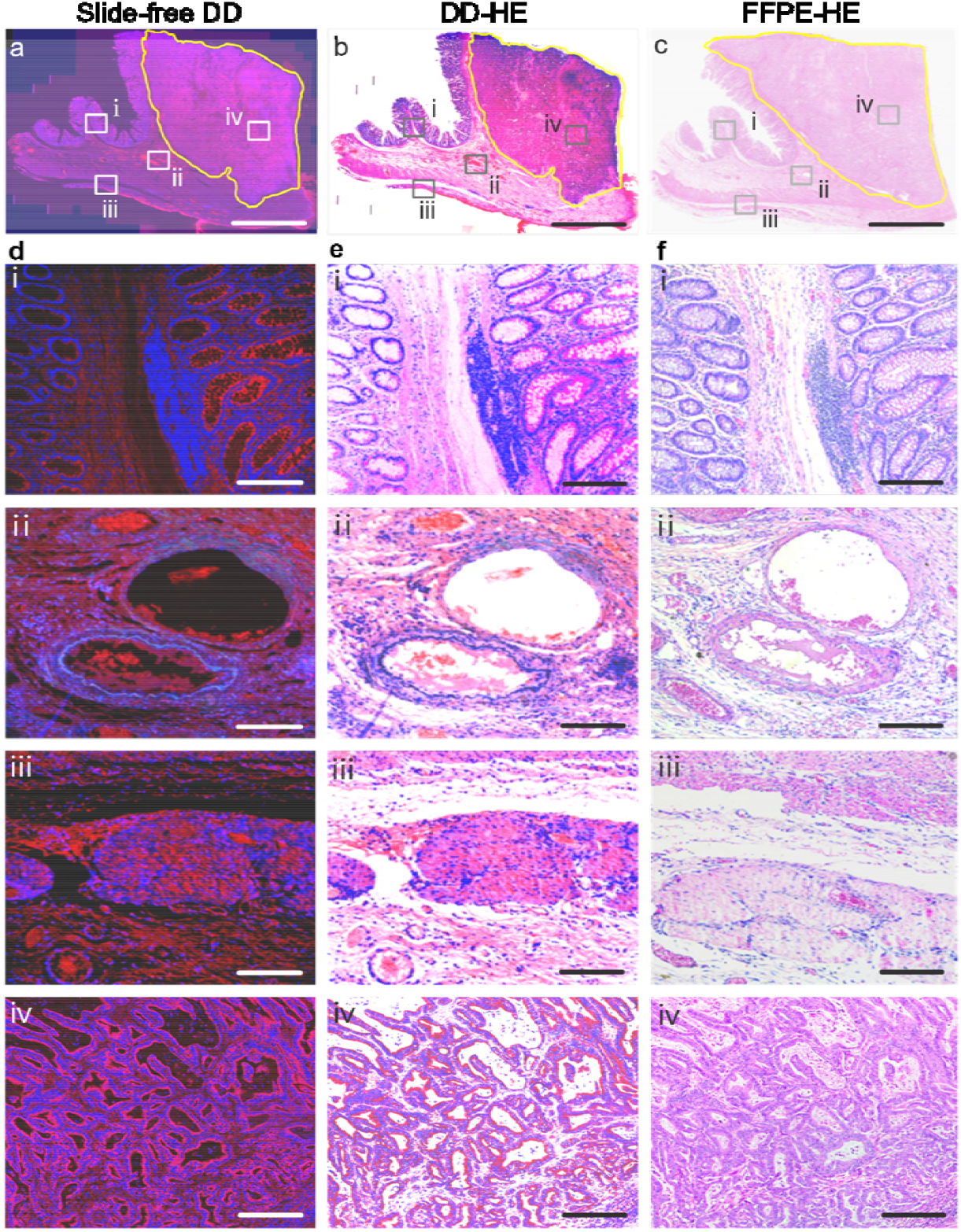
Comparison of key diagnostic morphological structures in human colon cancer tissue. **a, d**, Fluorescence images of colon tissue using the slide-free FLASH-Path method (DD image). **b, e**, Transformation of fluorescence images (**a, d**) to bright-field H&E-like images (DD-HE image). **c, f**, H&E images from standard H&E histopathology of the same tissue sample used in (**a**) (FFPE-HE image). All three image modalities clearly displa similar pathological structural features, such as normal colonic mucosal glands and interstitial lymphocytes (**i**), capillaries with red blood cells (**ii**), smooth muscle and adventitia (**iii**), and adenocarcinoma tissue (**iv**). Scale bars: (**a**-**c**) 5 mm; (i-**iv**) 200 μm.

After comparing DD-HE and standard FFPE-HE images from both image processing and pathologic diagnosis, we found that many evaluation indexes (for example, clarity, contrast, color fidelity, and noise control) ^30^ of DD-HE exceed or are equivalent to standard FFPE-HE images. Note that: (1) Good clarity means it is easy to visualize different subcellular structures (such as nucleus, cytoplasm) and intercellular matrix, as well as tissue levels and boundaries; (2) Good contrast is determined by bright blue for nucleus and well-defined red for cell membranes, and it easy to differentiate tissue layers and cell types; (3) High color fidelity is related to high accuracy and consistency of tissue and cell colors across different field of view (FOV); and (4) Good noise control means no additional signals or artifacts. In the DD-HE image shown in Fig. 2b, we can see many necessary structures for pathologic diagnosis, such as the mucosal layer and lymphoid tissue of human colon cancer tissue (Fig. 2b-i), the vascular lumen and red blood cells in the submucosal layer (Fig. 2b-ii), the smooth muscle layer and outer membrane (Fig. 2b-iii), and the area of the colon adenocarcinoma mass (in the yellow circle, see Fig.2b-□). We compare the DD-HE image (Fig. 2b) with the corresponding FFPE-HE image (Fig. 2c), and can’t see significant morphological differences which would affect pathologic diagnosis. Therefore, the high-quality slide-free pathology images from FLASH-Path are sufficient to ensure accurate identification of normal and heterogeneous tissue structures.

### 3. Comparative Imaging of DD staining and H&E Staining Reveals Identical Subcellular Architecture in the Same Tissue Section

Images of cytological structures (especially a detailed visualization of nuclei) are central to tumor pathology diagnosis ^31^. In order to strictly compare the imaging performance between FLASH-Path and conventional FFPE-HE, we selected a FFPE-sectioned colorectal tissues to perform DD staining and fluorescence imaging (see Fig. 3a), and then converted it to DD-HE image (see Fig. 3b). The same section was also stained with standard Hematoxylin & Eosin histopathology protocol and imaged using bright-field microscopy (Fig. 3c). We performed co-localization analysis on the extracted nucleus structures (excluded fatty regions) in the same FOV. We set the color channels as Red (DD), Green (DD-HE) and Blue (FFPE-HE), respectively, and obtained a fused image which was further used to perform co-localization analysis (see Fig. 3d). The structures in Fig. 3d were completely overlapped, as seen by the white color. From the magnified areas (Fig. 3e and Fig. 3f), almost all nuclei are shown in white, with some tiny purplish-red areas demonstrating that DD and DD-HE imaging are able to present more cellular structures than FFPE-HE.

**Fig. 3.**
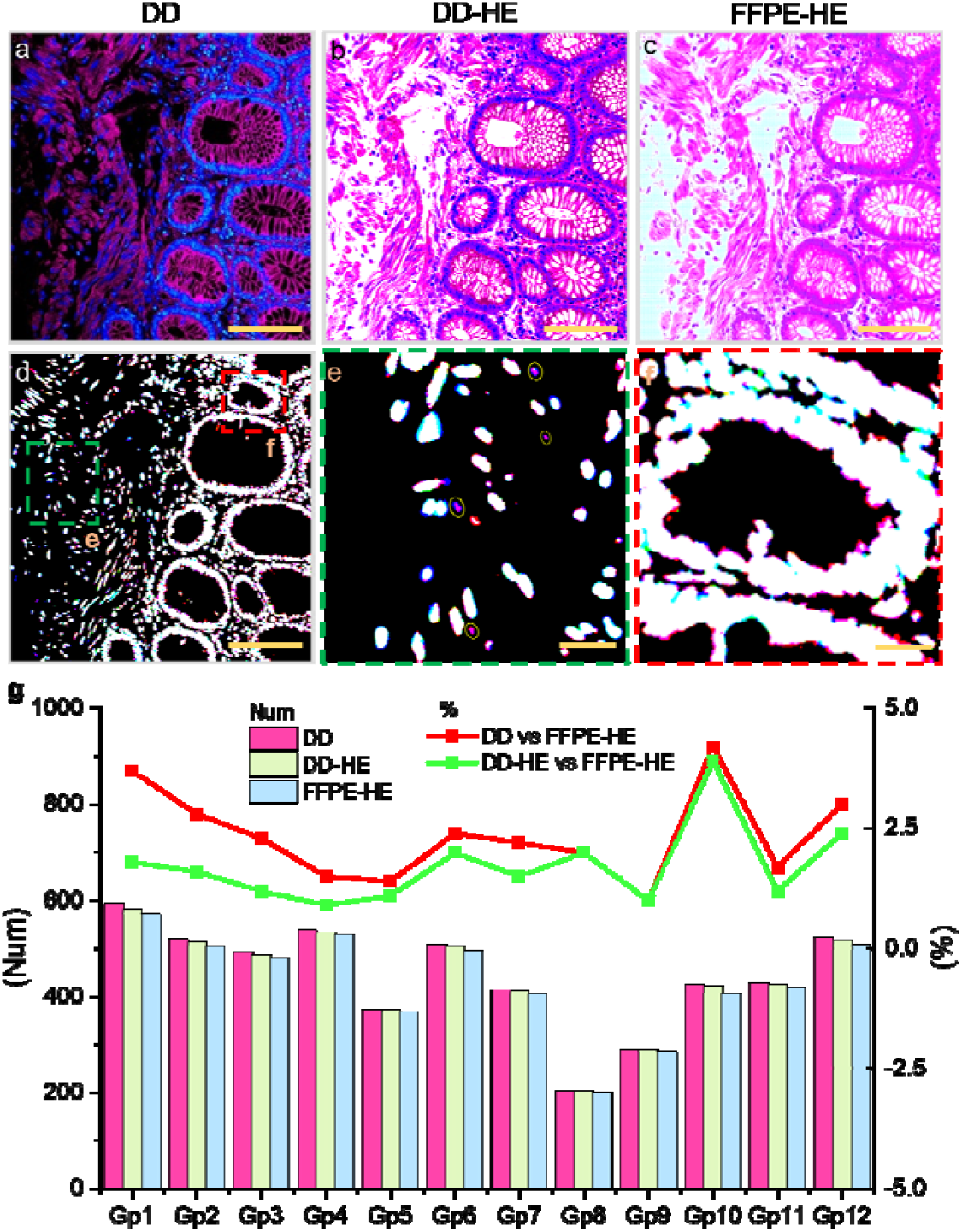
Imaging the same tissue slice using FLASH-Path and H&E. **a**, DD image. **b**, DD-HE image. **c**, FFPE-HE image. **d**, Fused images with extracted cell nucleus. The images from DD, DD-HE and FFPE-HE were set t red(R), green(G) and blue(B) color channels, respectively, and then fused into RGB image. **e-f**, Magnified views of the boxed regions in (d). **g**, Statistics (bar graphs) and differences (line graphs) of the number of detected nuclei in 12 random groups (abbreviated as Gp) of corresponding ROIs (1000 × 1000 pixels each) in (a-c). Se Supplementary Fig. S5 and Table S1 for the counted nuclei values. Scale bars: (**a-d)** 500 μm, (**e-f)** 50 μm.

In order to further quantify the differences among the three images, we randomly selected 12 sets of ROI images (1000×1000 pixels each) in Fig. 3a-3c (see supplementary Fig. S5 for more details), and counted the number of nuclei (see supplementary Table S1). As shown in Fig. 3g, we found that the number of counted nuclei varies in less than 5%, and the number of nuclei counted in the DD and DD-HE images are both higher than that in FFPE-HE. We checked again into the small structures in Fig. 3d and speculated that the DAPI dye used in DD staining might have stained some nuclear fragments or extra-nuclear DNA molecules ^32^, and these structures were not recognized in the FFPE-HE images because of low contrast. The superior nuclei imaging from DD staining not only verifies the clinical accessibility of the new technique, but also proves the artifact-free nature of the color conversion from DD image to DD-HE image.

### 4. Imaging Lipid-rich Tissues with FLASH-Path

The application of conventional frozen section pathology is limited not only by the ice crystal artifacts, but also by the fact that lipids have low freezing point and are easy to dissolve in organic solvents. Therefore, lipids in tissue samples must be removed as much as possible before preparing frozen sections ^33^. In contrast, the FLASH-Path technique developed in this paper eliminates the need for tissue sectioning and various chemical solvent treatments. The selection of lipophilic DiD dyes further increases the ability to recognize structures with lipids.

To verify that fat cells can be imaged by DD staining, we performed rapid slide-free DD staining and FLASH-Path imaging on the cross section of mouse colon tissue (see Fig. 4a, with a diameter of 2-3 mm), and the image was converted into DD-HE (see Fig. 4b). After FLASH-Path imaging, FFPE-HE imaging was performed in the same tissue according to the standard protocols, and the result is shown in Fig. 4c. Comparing DD-HE (Fig. 4b) with FFPE-HE (Fig. 4c), we can see the tissue structures of mouse colon, especially the important structures for pathologic diagnostics (intestinal mucosal epithelial tissue, cell nuclei, and cytoplasm) are well presented, with basically the same clarity and contrast (as seen in Fig. 4e-v and 4e-vi). Notably, a closer look at the extra-intestinal regions with adipocytes (see Fig. 4d-□ and 4d-iii) reveals that the distribution of nuclei remains the same in both DD-HE and FFPE-HE images; however, the DD-HE image provides a better visualization on the cytoplasm and cell membrane structures. Therefore, the FLASH-Path technique can be used for high-quality imaging of samples containing adipose tissues.

**Fig. 4.**
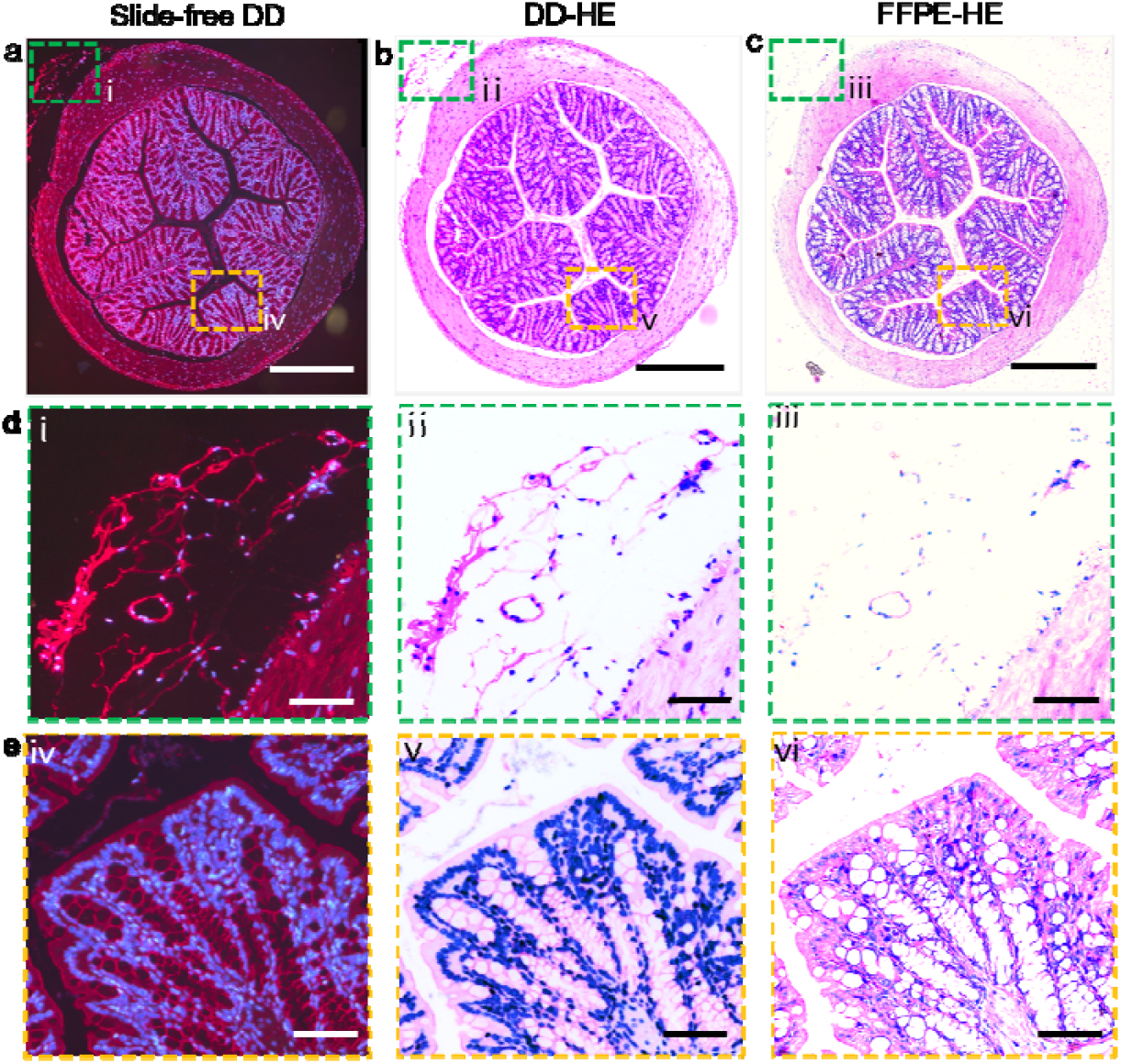
Comparative imaging of adipocytes using FLASH-Path versus conventional FFPE-HE in lipid-rich tissues. **a, b and c** correspond to Slide-free DD, DD-HE and FFPE-HE images. **d(**i**) - (iii)** correspond to magnifie views of the adipose tissue regions in **a-c** (green boxes); **e(iv) - (vi)** correspond to magnified views of the normal colorectal mucosal tissue in **a-c** (yellow boxes). Mouse colon tissue was used. Scale bars: (**a-c**) 1 mm, (**d, e**) 200 μm.

### 5. Imaging Large Tissue Sample with FLASH-Path

Due to the limitations of embedding cassette size, slide area, and the movement range of the normal microscope stage, the Pathology Technical Guidelines recommend that the optimal tissue block for pathology sampling is 15×15 mm in size and 3-5 mm in thickness ^34^. However, in actual clinical scenarios, the size of most surgical specimens exceeds the recommended values. Therefore, pathologists usually cut the large-volume samples into standardized small pieces, which not only increases the workload and diagnostic difficulty, but also destroys the integrity of the samples. In contrast, FLASH-Path enables direct large-area imaging of fresh tissue specimens, eliminating the workflow of sample fragmentation and multi-section imaging. And, the imaging coverage of FLASH-Path is no longer constrained by cassette dimensions but determined by the stage movement range. As a representative example, we were able to perform slide-free imaging (Fig. 5a) and color conversion (Fig. 5b) for samples with an area of more than 35×45 mm.

**Fig. 5.**
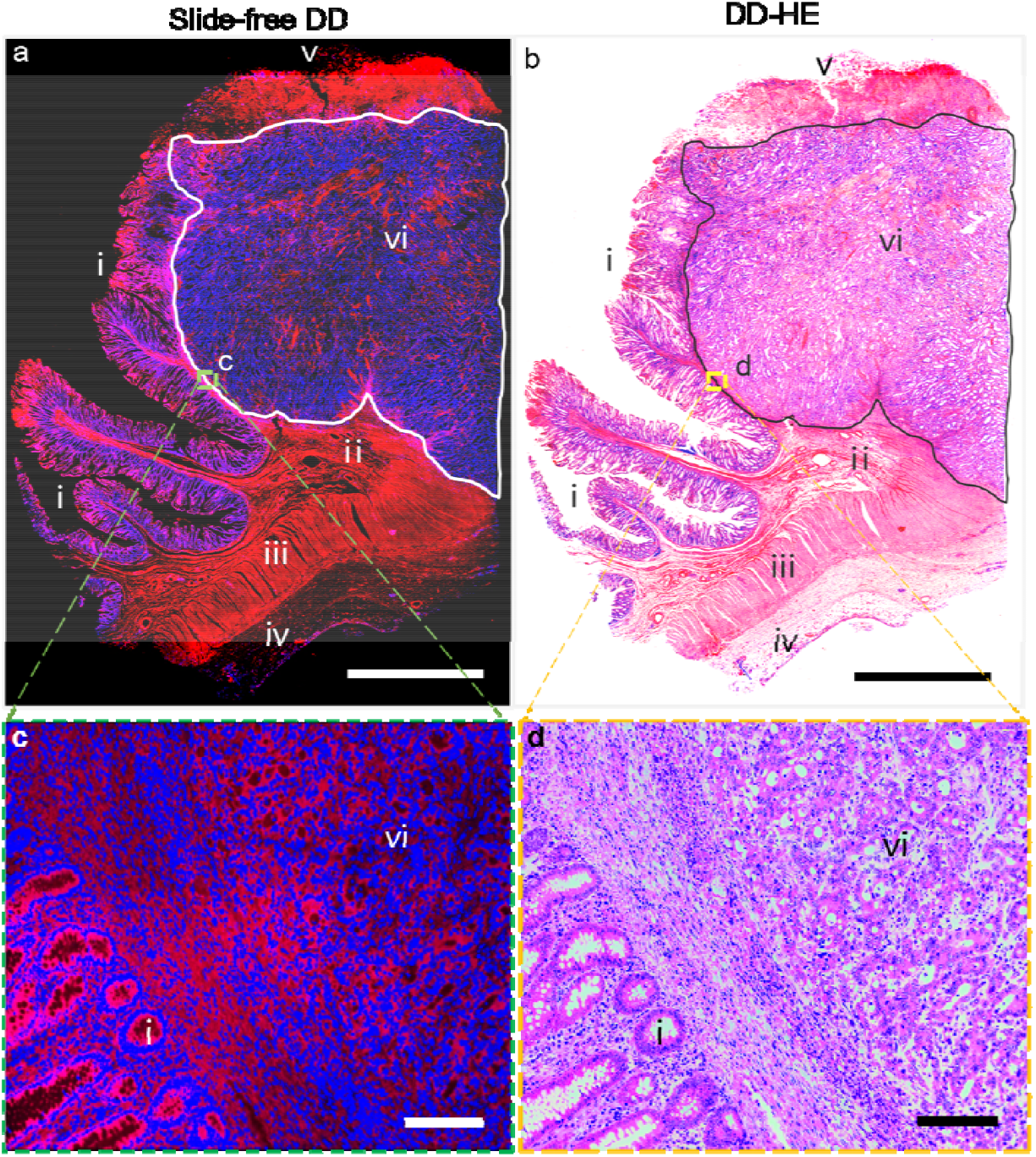
Slide-free imaging of large sample using FLASH-Path. **a**, DD image. **b**, DD-HE image. **c-d** Magnifie views of the boxed area in (**a-b**). Scale bars: (**a, b**) 1 cm, (**c, d**) 200 μm.

It is worthy to point out that FLASH-Path not only significantly increases the imaging area, but also completely maintains good image quality across the entire imaging area. As seen in Fig. 5b and 5d, the key tissue structures for pathologic diagnostic ^4^ can be clearly rendered in the DD-HE images, including the normal colorectal mucosa (□), submucosal layer (□), smooth muscle layer (□), epithelial layer and extra-intestinal fat (□), necrotic tissues on the surface of the mass (□), and colon adenocarcinoma mass (vi). From a sense of image resolution and contrast, we enlarged the junction of adenocarcinoma mass and normal colorectal tissues (the yellow box in Fig. 5b), and are able to observe the normal colorectal glands with orderly cellular arrangement (□), the cancerous area with disorganized cellular arrangement and obvious heterogeneity (vi), and the tumor boundary. These results verify that the rapid, slide-free FLASH-Path technique is able to image both small samples (Fig. 4) and large samples (Fig. 5) while still maintaining superior image quality (comparable or even better than conventional FFPE-HE), thus providing better support for pathology diagnosis.

## Discussion

The FLASH-Path technique overcomes fundamental limitations of conventional mechanical sectioning and optical sectioning approaches through an innovative thin-layer tissue staining paradigm. This slide-free platform enables rapid histopathological evaluation of centimeter-scale clinical specimens within 10 minutes - a critical time window for intraoperative decision-making in emergency surgical settings.

The FLASH-Path platform enables high-fidelity visualization of ultra-thin tissue sections (≤ 10 μm thickness) through an optimized dual-channel fluorescence strategy combining lipophilic membrane-selective DiD and nucleic acid-targeting DAPI staining. This histological imaging advancement is achieved through integrated large-area mosaic scanning protocols and extended depth-of-field (EDF) optical sectioning, which synergistically address resolution-depth tradeoffs inherent in conventional whole-slide imaging systems ^35^. High-fidelity H&E-like images are generated through physics-driven spectral decomposition-based color transformation, where each pixel’s origin is traceably mapped to distinct histological substructures. This AI-independent approach maintains intrinsic biological interpretability by preserving direct structure-color correlations through optical property quantification, effectively bypassing the latent space uncertainties inherent in AI-mediated image synthesis. The methodology demonstrates superior clinical relevance as evidenced by pathologists’ validation of diagnostic equivalence to gold-standard H&E staining, while fundamentally eliminating hidden risks associated with AI-generated structures. Specifically, cross-species experimental validation using mouse colon specimens and human CRC tissue samples demonstrated the FLASH-Path technique’s comparable or superior performance relative to FFPE-H&E histopathology in imaging speed, sample area, and image quality. The quantifiable technical superiority of FLASH-Path was also demonstrated in the presentation of adipose tissue and lymphatic structures, which significantly outperforms the gold-standard frozen section histopathology.

Compared with the gold-standard FFPE-HE and frozen section histopathology techniques (see Fig. 6), the FLASH-Path system overcomes the problems of complex sample preparation and freezing artifacts, while enabling large-sample imaging (≥35×45 mm) within 10 minutes, surpassing the sample size limitations in mechanical sectioning and enhancing intraoperative pathology potentials. Relative to slide-free technologies (e.g., MUSE, UV-PAM), FLASH-Path replaces optical thin-layer excitation with controlled thin-layer staining. Built upon clinical-accessible fluorescence microscopy, FLASH-Path addresses the operation difficulty and cost due to the complex optical systems, and substitutes virtual H&E staining with traceable color conversion, improving interpretability and medical evidence. Thus, FLASH-Path provides superior clinical accessibility for rapid pathology diagnostics compared to traditional and emerging technologies.

**Fig. 6.**
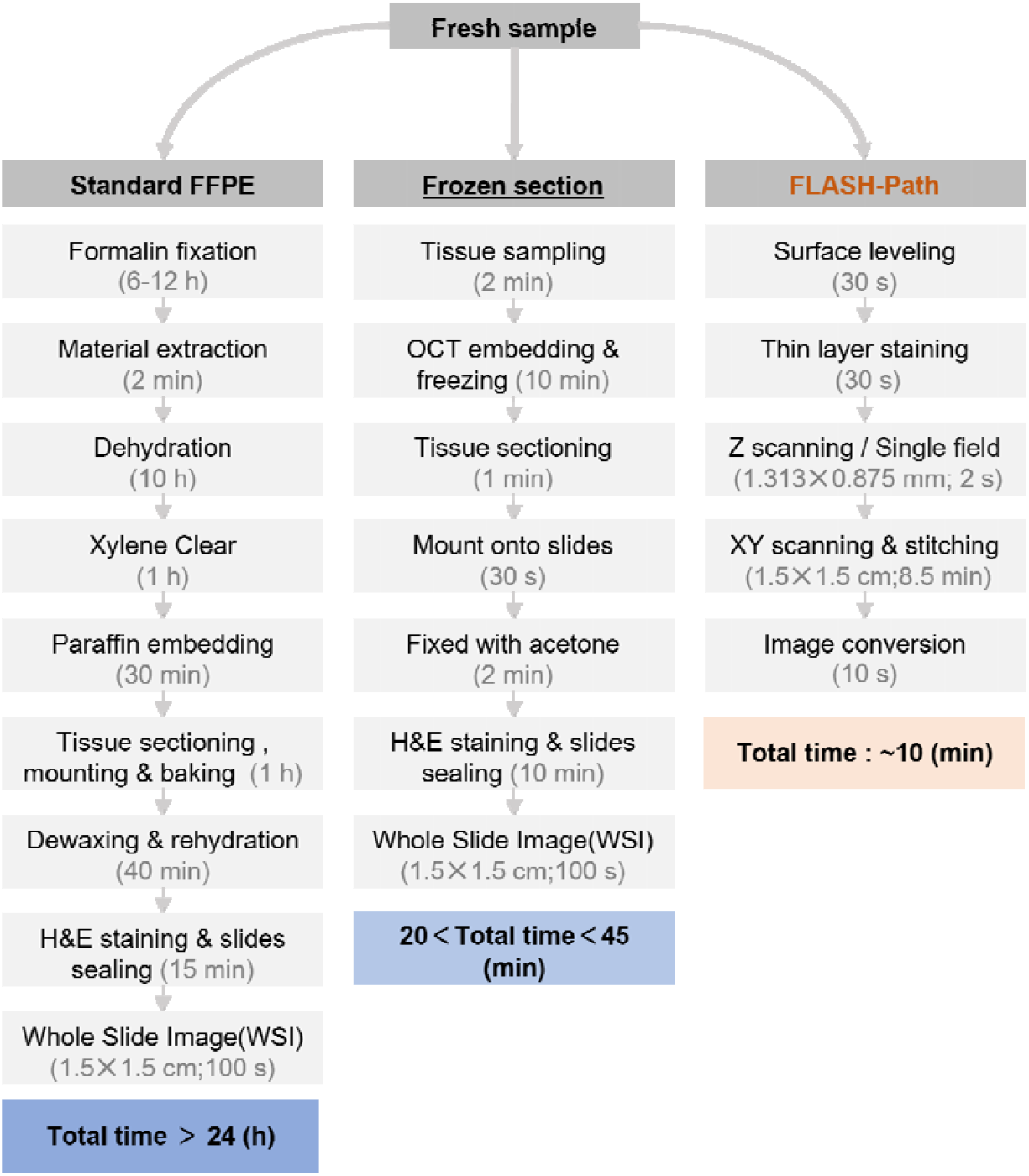
Comparison of FLASH-Path imaging workflow with the gold-standard FFPE-HE and frozen section histopathology.

The innovation of this study lies in the combination of rapid thin-layer staining, large-area mosaic scanning and extended depth-of-field (EDF) imaging, which realizes slide-free, rapid and high-quality imaging of diverse tissue samples. Its clinical translational value is significant, not only can reduce the equipment acquisition and operation cost of hospitals, but also shorten the technical training time by simplifying the operation process, which has high generalizability. In addition, the FLASH-Path system can also support remote transmission and consultation through networked devices, further enhancing its potential for clinical application. The technology provides an efficient, cost-effective and promising solution for intraoperative pathologic diagnosis, which is especially valuable in surgeries such as mid-low rectal cancer that require precise assessment of the margins. Particularly noteworthy is the fluorescent image of membrane red and nuclear blue, which is equivalent to the fluorescent version of the H&E staining effect, and can be directly used in emergency diagnostic scenarios even without color conversion.

Despite the system’s excellent performance in rapid pathologic diagnosis, there are still some limitations. First, the system is currently focused on staining and imaging cell membranes and nuclei ^36^, and further labeling of specific biomarkers is required for certain lesion cells that need to be typed and graded ^37^. Secondly, the assessment of the nuclear-to-cytoplasmic ratio, which is usually used to determine the malignancy of a tumor, can only be identified morphologically after systematic imaging which has to be performed by a pathologist based on specialized training. In future studies we will add the possibility of automatic identification of tumor cells based on the staining dominance of cell membranes and nuclei. Third, the imaging speed of the system is limited by the frame rate of the detector (currently 14 frames per second, FPS) and the imaging speed can be further improved by using cameras with higher sensitivity and frame rate in the future. In addition, the imaging speed of the system depends, in part, on the intelligent control of the XYZ automatic stage, and a good hardware platform and control system are also the key guarantee of imaging speed, but this may add to the cost and difficulty of operation. In this paper, in addition to using colorectal cancer samples as a performance validation subject, we also performed imaging experiments on a variety of other organ tissues, all of which showed excellent results comparable to those of colorectal cancer samples in slide-free imaging. Unfortunately, a high image background was found when imaging non-decalcified treated bone tissues, and we hypothesize that the background color is the autofluorescence of minerals such as calcium phosphate ^38^, and we look forward to resolving this issue in subsequent studies to make the FLASH-Path system more broadly generalizable.

In summary, the FLASH-Path technology has a wide range of application potentials in the field of intraoperative pathology diagnosis. On one side, the imaging quality and interpretability of this technology make it more acceptable to doctors. On the other side, the low cost and high generalizability of this technology make it promising to spread its use in hospitals at all levels, especially in resource-limited areas, and to reduce the healthcare burden of patients. Furthermore, by the upgrading with immunofluorescence staining, AI-empowered diagnostic pathology ^39,40^ and other technologies, FLASH-Path can also be applied in tumor grading and other fields to further realize more clinical value.

## Methods

### 1. Thin-layer fluorescence staining

In the experiments, we first performed rapid surface smoothing and decontamination of the samples to be tested. Subsequently, we configured the DiD staining reagent (Beyotime C1039-10mg, diluted to 10 μm/L) with a 1% mass fraction of aqueous hyaluronic acid as the solvent. Hyaluronic acid molecules can absorb hundreds of times their own weight of water molecule and form a gel film covering the tissue surface ^41^, thus realizing the uniform release of probe molecules in the covered area and achieving the purpose of controlled permeation of the solution. The specific labeling process is as follows: (A) Sample pretreatment: rinse the flattened sample 3 times with 1×PBS buffer (PBS, Sigma-Aldrich P4417, pH 7.2-7.6, isotonic); gently absorb the water on the surface of the sample with a filter paper. b. DiD-staining: DiD (10 μm/L) configured with 1% aqueous hyaluronic acid was used for surface staining of the pretreated samples. (B) DiD-staining: the surface of pretreated samples was stained with DiD (10 μm/L) in 1% aqueous hyaluronic acid solution for 20-30 seconds; residual DiD reagent on the surface of the samples was washed away with PBS. (C) DAPI staining: the samples were stained by infiltration with DAPI (Sigma-Aldrich MBD0015, 5 mg/ml) solution for 30 seconds; the samples were rinsed with PBS for three times to wash away unbound DAPI reagent on the surface. (D) Sealing solution: the samples were stained by infiltration with the appropriate amount of anti-fluorescent quenching sealer (SlowFade™ Glass, ThermoFisher, Sterilizer) Glass, ThermoFisher, S36917-5X2ML) onto the imaging surface of the tissue; cover the surface of the tissue with a 0.17 mm quartz coverslip. (E) Precautions: The tissue should be secured in a Petri dish with the test surface facing upward; the base of the Petri dish is made of a molded soft body to support the back of the sample to ensure that the test surface of the sample is at the same level as the stage of the orthoptic microscope, which helps to reduce the depth of field and expand the imaging surface. This helps to minimize the difficulty of EDF imaging. Once the above process is completed, the sample can be scanned and imaged.

In order to verify the effect of thin-layer staining, we performed three-dimensional imaging of samples with different lengths of staining time using a confocal microscope (Olympus FV3000) at 20× magnification, and the experimental results are shown in supplementary Fig. S3. The infiltration depth of the probe staining on the samples increased gradually with the time of the staining treatment, indicating that controlling the staining time can effectively control the infiltration depth of the dye (see supplementary Fig. S3b and Fig. S3c,). So, the infiltration depth in the direction of the thickness of the sample increases sharply with the increase of the dyeing time, and after reaching a certain depth, it decreases exponentially (see Fig. 1b). The reason for this phenomenon is supposed to be the rapid diffusion of molecules in the direction of low concentration before removing the osmotic pressure of the solution on the surface of the sample, and after removing the osmotic pressure of the solution on the surface of the sample, the diffusion speed of molecules decreases rapidly, and the process of combining with the cell membrane on the diffusion path further consumes the molecules, which makes the depth of molecular infiltration decrease rapidly ^42^.

### 2. Human CRC preparation

The human CRC samples in this study were obtained from residual colon cancer tissue preserved in 4% neutral buffered formalin after surgical resection. Upon receipt of these samples, a scalpel was first used to excise a large block of tissue from the interface between the lesion and normal colonic tissue. Next, interfering materials such as cauterized tissue and blood clots were removed from the imaging surface, and excessive tissue surface irregularities were corrected using a razor blade or other tools. Finally, surface debris was removed, and the tissue was rinsed with PBS buffer to eliminate free impurities. Fluorescent labeling was performed according to the previously described thin-layer staining protocol. After the slide-free imaging, the tissue was processed using standard FFPE-HE histological techniques for sectioning and staining, followed by whole-slide bright-field microscopy to obtain comparative H&E images.

The human colorectal samples used in this study were provided by the Department of Pathology, Beijing Anzhen Nanchong Hospital of Capital Medical University & Nanchong Central Hospital. The Medical Ethics Review Committee of Beijing Anzhen Nanchong Hospital of Capital Medical University & Nanchong Central Hospital granted ethical approval for this research.

### 3. Mouse colon samples

The mouse colon paraffin sections used in this study were obtained from KM mice purchased from Wuhan Servicebio Technology Co., Ltd., China. The tissue samples underwent three deparaffinization steps before being subjected to DD staining and imaging, following the previously described staining protocol. After imaging, the residual dye was partially removed using a gradient ethanol wash, and standard H&E staining was performed for imaging, allowing for point-by-point structural comparison with the DD images of the same sample.

### 4. Image acquisition

The imaging system used in this study was built upon the main body of an Olympus microscope (BXFM, Olympus) configured with two objective lenses, 10× (UPLFLN10XP, Olympus) and 4× (PLN4X, Olympus). The system used an XY motorized stages (LAB-XY-50-50-B, LAB Motion Systems) to control the lateral movement of the sample, a stepper motor (PKP546N18A2, Oriental Motor) to control the axial movement of the sample along the objective lens, and a color camera (FL20, Tucsen Photonics) for image detection. The current system spatially combines a 405 nm laser and a 640 nm laser using a dichroic mirror (ZT 488/532/633/830/1064 rpc-UF1, Chroma), supplemented by lens systems with focal lengths of 100 mm (AC254-100-A-ML, Thorlabs) and 75 mm (AC254-75-A-ML, Thorlabs) for beam modulation, enabling full-field dual-channel excitation of DAPI and DiD dyes. To ensure high quality and accuracy of the imaging signals, the system is equipped with an excitation filter (FF01-390/482/532/640-25, Semrock) for laser purification, a dichroic mirror (ZT405/488/532/640 RPC-XT, Chroma), and an emission filter (ZET405/488/532/640 nm, Chroma) to separate and filter the fluorescence signals emitted from the sample. When the system is configured with a 10× objective lens, the imaging FOV is 1313.3 μm × 875.5 μm, and the imaging depth of field is about 12 μm. Therefore, when imaging biological samples, the sample lateral travelling pace is 85% of the imaging FOV, the axial travelling pace is the depth of field of the objective lens, and the number of travelling steps can be set by yourself according to the sample. For example, if the undulation range of the sample surface is 50 μm, five images should be acquired in the axial direction of the same FOV. In order to fuse the images with different focal positions in the axial direction together to give an EDF microscopic image, this paper uses the fact of rapid decay of signal intensity with the out-of-focus distance, and implements a real-time DOF fusion algorithm based on the extreme value projection (see Fig. 1c and supplementary Fig. S2). According to the equipment parameter in the imaging system, it can be estimated that imaging a sample with an area of 15×15 mm and a surface undulation of 50 μm requires 1020 FOVs at a magnification of 10×, and the time consumed is about 8 minutes. This process takes much less time than the H&E preparation process and is faster than the frozen section imaging process, which can fully meet the rapid imaging needs for intraoperative pathology diagnosis.

### 5. Image processing

Although the DD images acquired by the FLASH-Path system already encompass all the pathological features, the image reading habits of pathologists were considered in this paper by converting the fluorescence-mode images (DD images) into bright-field H&E-like images (DD-HE images). In practice, we first split a DD image into red, green, and blue channels. Then, using Equation (1.1), we performed image calculations to obtain a H&E-like image, referred to as the DD-HE image. This image transformation process involves simple addition and subtraction operations (See Supplementary Fig. S4 for details), which can be implemented using software platforms such as ImageJ or MATLAB.

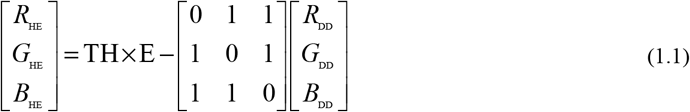

Where *R*_HE_*∼B*_HE_ represent the three channels of the DD-HE image, TH is a constant corresponding to the maximum grayscale value for the image bit depth (e.g., 255 for an 8-bit image, 65,535 for a 16-bit image), E is the identity matrix, and *R*_*DD*_*∼B*_*DD*_ are the three sub-channel images of the DD fluorescence image.

## Author Contributions

Z. L. H., Y. J. Z. and X. G. G. designed and led the experimental project. Y. J. Z., X. G. C. and Z. L. H. designed and fabricated the imaging systems used in the study. X. G. G. and Y. W. prepared the samples. Y. J. Z. and X. G. G. performed the sample staining experiments. X. G. G. performed confocal microscopy experiments. X. G. G. and X. G. C. performed image acquisition. F. H. Q. and X. G. G. performed localization analysis of the nucleus. Y. J. Z., X. G. C. and Z. X. W. performed data processing. Y. L. L. and X. G. G. performed documentation and translation. All authors were involved in data analysis and image processing.

## Acknowledgments

We thank all the other members of Digital Therapeutics and Optical Microscopy (Digi-TOM) group for their technical support. Funding for this work was provided by National Natural Science Foundation of China (82260368), National Key Research and Development Program of China (2022YFC3400601), Innovational Fund for Scientific and Technological Personnel of Hainan Province (KJRC2023C03), Hainan Province Science and Technology Special Fund (ZDYF2022SHFZ126) and Start-up Fund from Hainan University (KYQD(ZR)-20077).

## Data availability

Codes and data underlying the results presented in this paper are not publicly available at this time but may be obtained from the authors upon reasonable request.

## Additional information

Supplementary information.

