## Supplementary Figures 1-6, Table 1 and Notes 1&2 for "Rapid slide-free histopathology via fluorescent layer-adaptive subcellular imaging: clinical-accessible platform for diverse tissues"

| **Table of contents** | | |
| --- | --- | --- |
| **Content** | **Description** | **Page** |
| **Figure S1** | Information for determining the excitation wavelength | 3 |
| **Figure S2** | Illustration of extended depth-of-field imaging and panoramic fluorescence imaging | 4 |
| **Figure S3** | Confocal microscopy images of DD staining at different durations for non-sectioned human colon tissues | 5 |
| **Figure S4** | Workflow for converting fluorescence microscopy images into DD-HE images | 6 |
| **Figure S5** | Images for counting nuclei numbers | 7 |
| **Figure S6** | An enlarged image showing the nuclei counting | 9 |
| **Table S1** | Statistical results of nuclei for each ROI in Supplementary Fig. S5 | 10 |
| **Note 1** | The fluorescence excitation wavelengths for the two dyes | 11 |
| **Note 2** | Tissue surface flattening | 12 |

**Supplementary Figure S1**

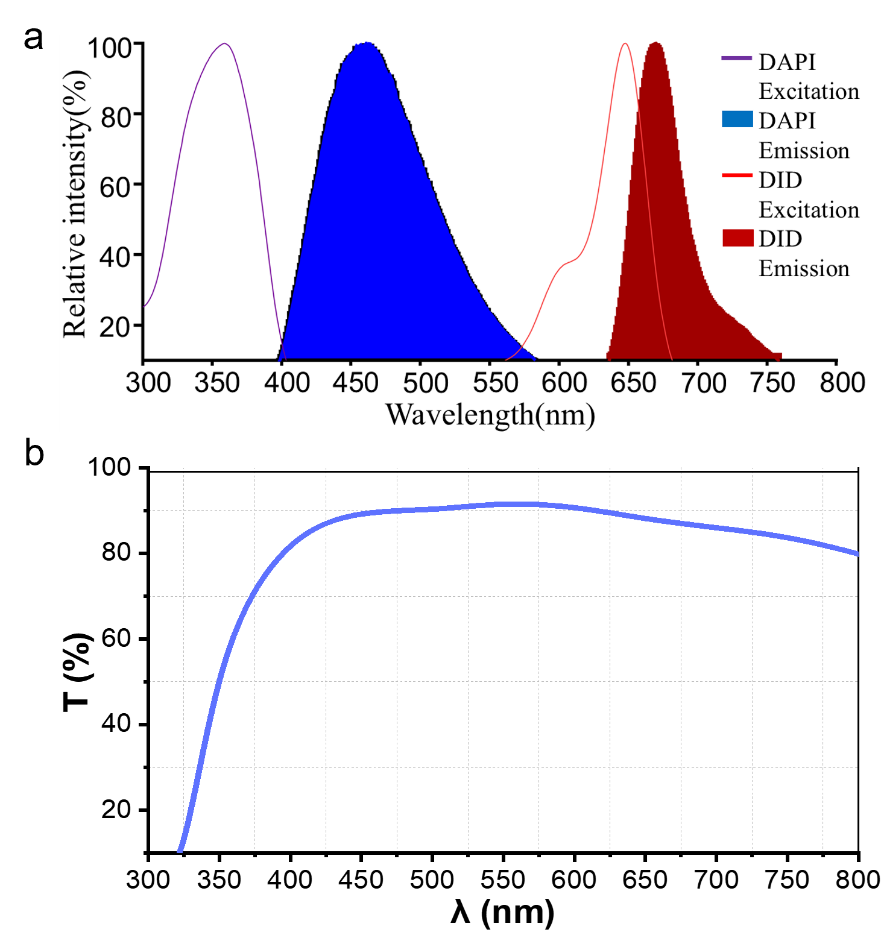

**Fig. S1 | Information for determining the excitation wavelength.** **a** Excitation and emission spectra of the nuclear probe DAPI and the cell membrane probe DiD. **b** Wavelength-dependent transmittance curve of a 10× objective (UPLFLN10XP, Olympus) mounted on a conventional fluorescence microscope.

**Supplementary Figure S2**

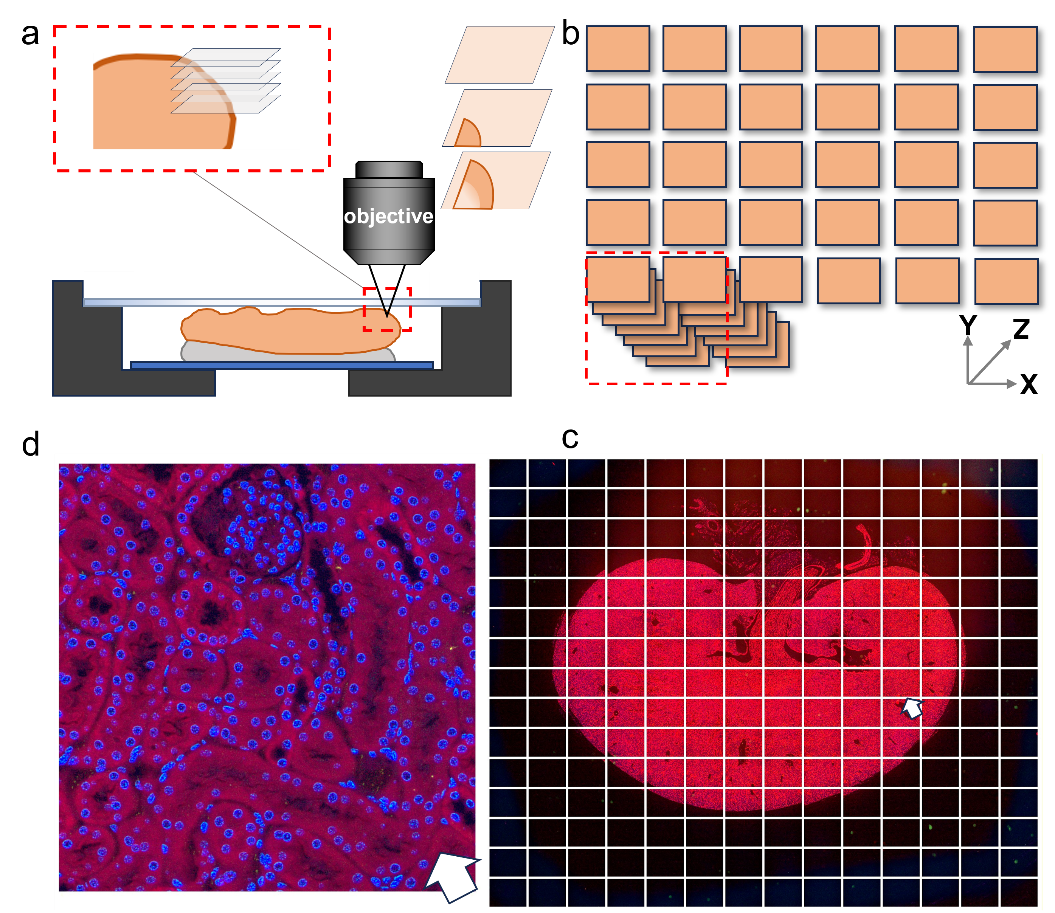

**Fig. S2 | Illustration of extended depth-of-field imaging and panoramic fluorescence imaging. a** Extended depth-of-field imaging. **b** Image stitching of multiple field-of-view images after extended depth-of-field imaging and image fusion. **c** Captured images in the image acquisition interface. **d** A magnified region from the image acquisition interface.

**Supplementary Figure S3**

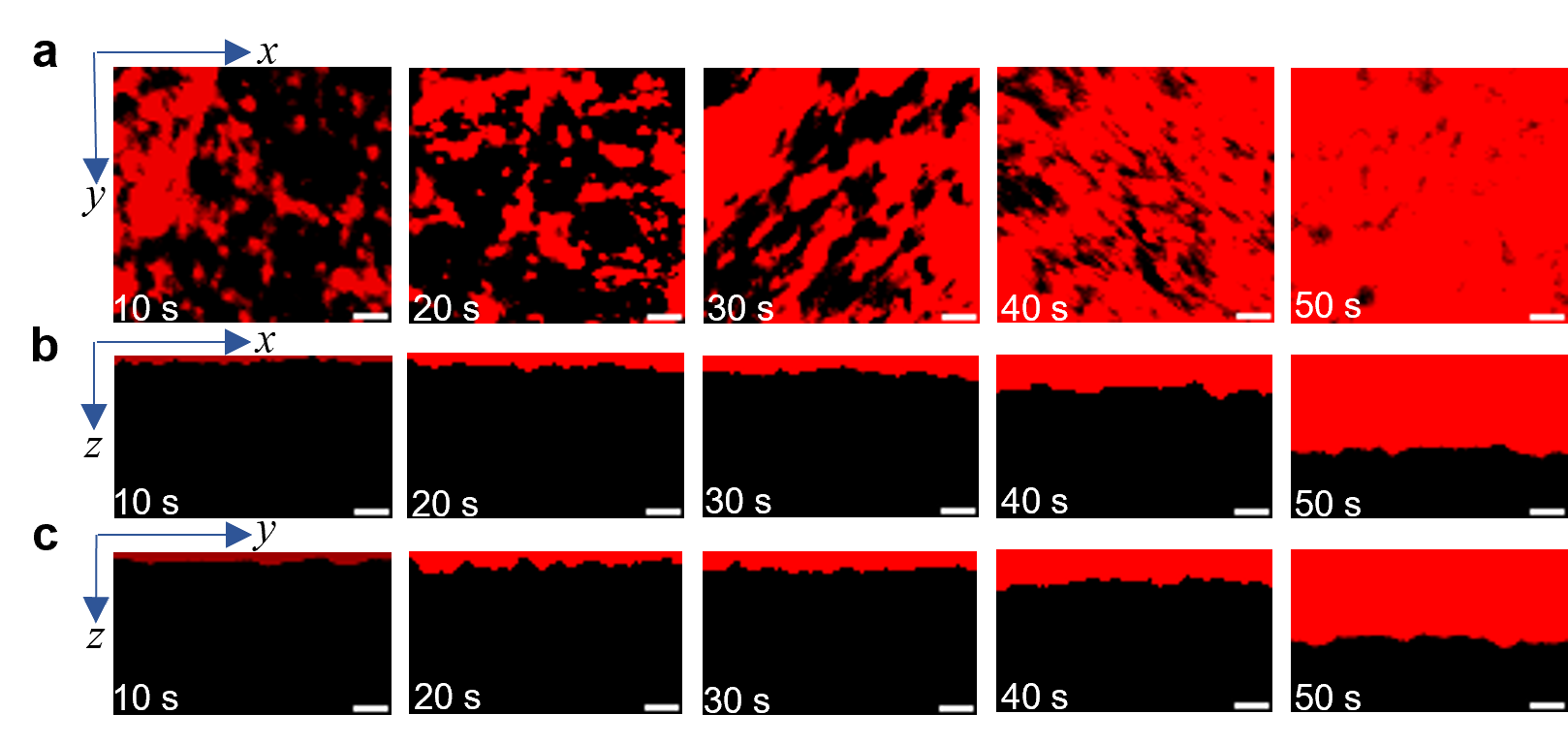

**Fig. S3 | Confocal microscopy images of DD staining at different durations for non-sectioned human colon tissues. a** Images in the XY plane. **b** Images in the XZ plane. **c** Images in the YZ plane. Scale bar: (**a-c)** 10 μm.

**Supplementary Figure S4**

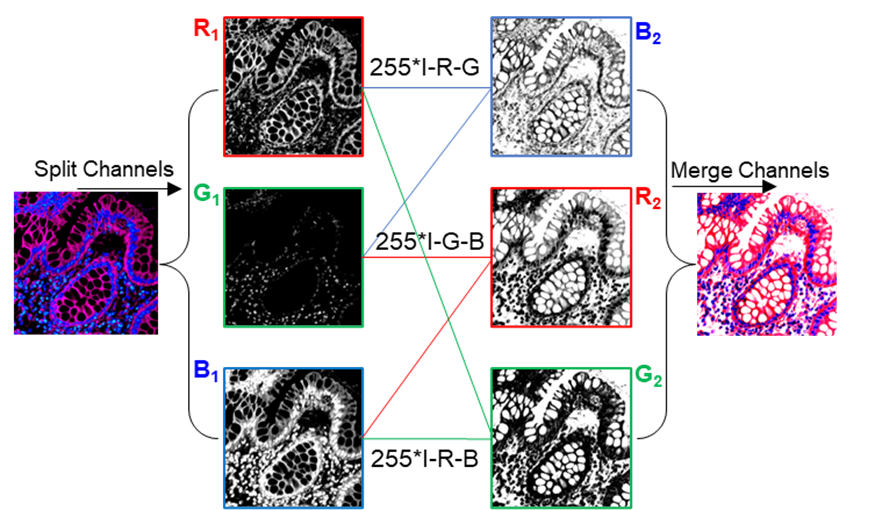

**Fig. S4 | Workflow for converting fluorescence microscopy images into DD-HE images.** The original fluorescence images stained with DAPI and DiD are decomposed into three color channels: R1 (Red), G1 (Green), and B1 (Blue). These color channels are converted into new color channels (R2, G2, and B2) based on the specific formulas shown inside the image. Channel merging and parameter adjustment are performed using the open-source software ImageJ to finally generate an H&E-like image (DD-HE) that resembles H&E staining.

**Supplementary Figure S5**

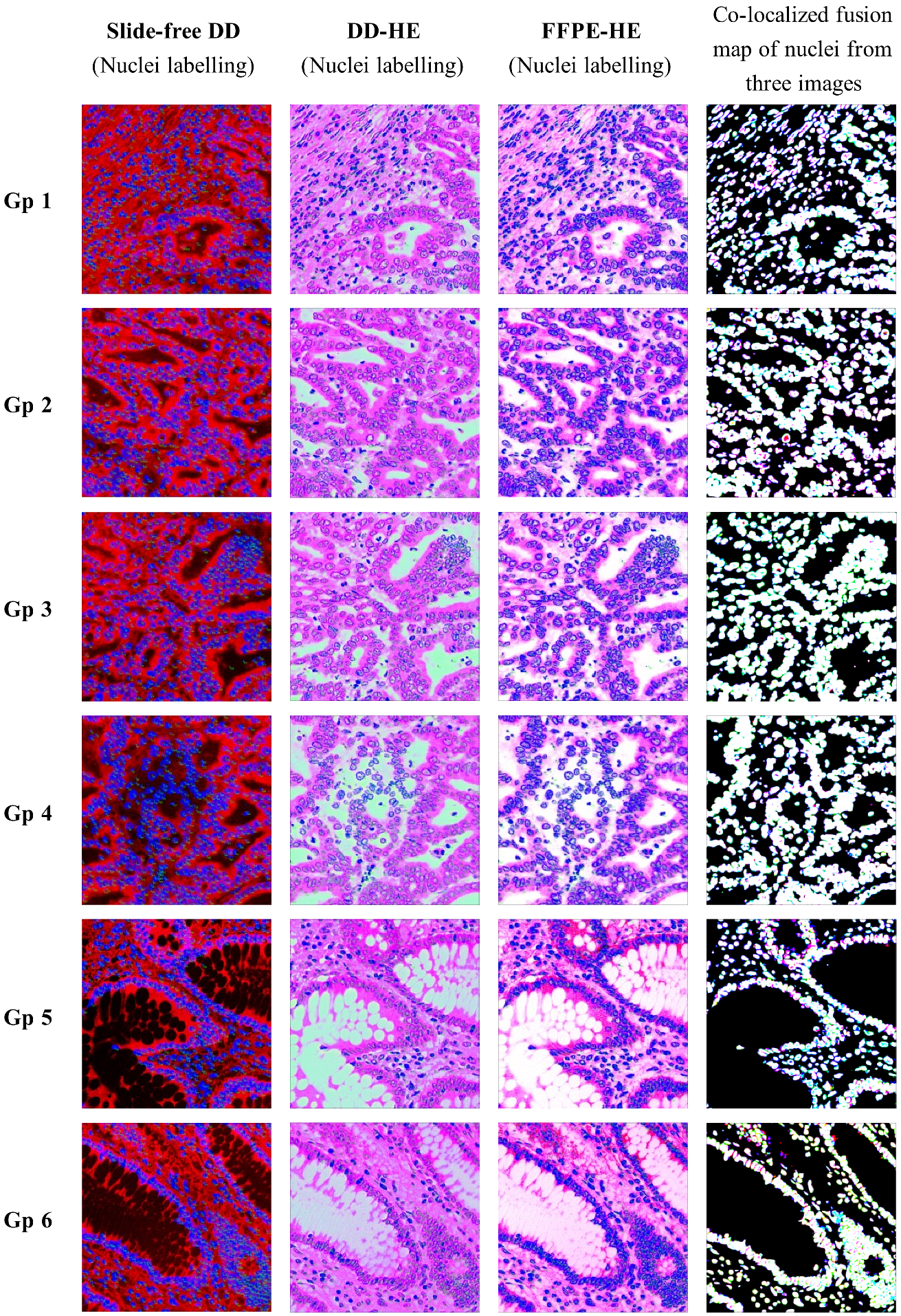

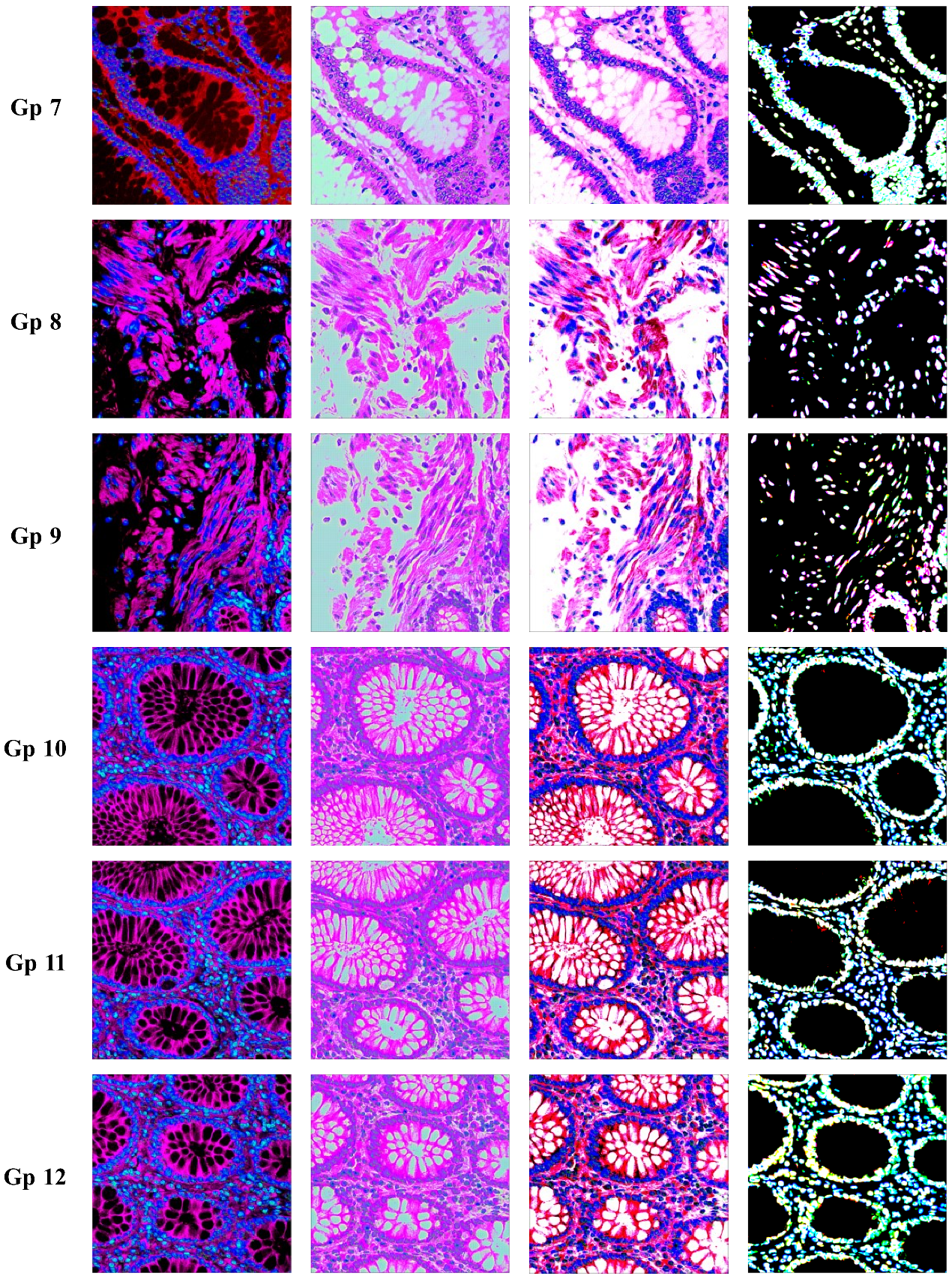

**Fig. S5 | Images for counting nuclei numbers.** Nuclei in the same tissue section were stained and imaged with both FLASH-Path and H&E histopathology. A total of 12 ROIs (120 μm ×120 μm, or 1000×1000 pixels) were selected for fusion and co-localization. The nuclei were set to red (DD), blue (FFPE-HE), and green (DD-HE). The overlapping nuclei were set to magenta (DD & FFPE-HE), yellow (DD & DD-HE), and cyan (FFPE-HE & DD-HE). The group numbers (**Gp**) are shown on the left.

**Supplementary Figure S6**

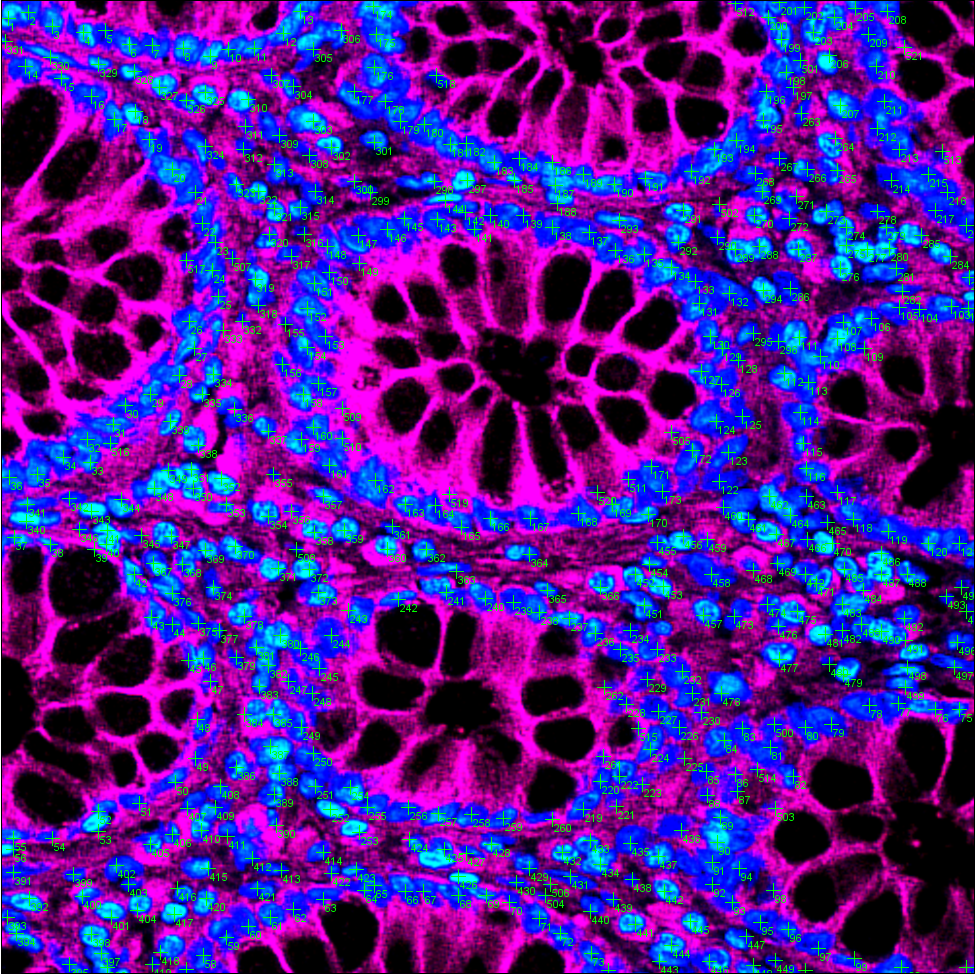

**Fig. S6 | A representative image showing the counted nuclei.** The image was enlarged from Supplementary Fig. S5 (Gp12).

**Supplementary Table S1:** Statistical results of the counted nuclei in Supplementary Fig. S5.

| **Group number** | **Nuclei number in the DD image** | **Nuclei number in the DD-HE image** | **Nuclei number in the FFPE-HE image** | **Nuclei number difference**  **between**  **DD and DD-HE** | **Nuclei number difference**  **between**  **DD-HE and FFPE-HE** |
| --- | --- | --- | --- | --- | --- |
| **Gp 1** | 592 | 581 | 571 | +21 | +10 |
| **Gp 2** | 519 | 513 | 505 | +14 | +8 |
| **Gp 3** | 492 | 487 | 481 | +11 | +6 |
| **Gp 4** | 537 | 534 | 529 | +8 | +5 |
| **Gp 5** | 373 | 372 | 368 | +5 | +4 |
| **Gp 6** | 507 | 505 | 495 | +12 | +10 |
| **Gp 7** | 414 | 411 | 405 | +9 | +6 |
| **Gp 8** | 203 | 203 | 199 | +4 | +4 |
| **Gp 9** | 289 | 289 | 286 | +3 | +3 |
| **Gp 10** | 423 | 422 | 406 | +17 | +16 |
| **Gp 11** | 426 | 424 | 419 | +7 | +5 |
| **Gp 12** | 521 | 518 | 506 | +15 | +12 |

**Supplementary Note 1. The fluorescence excitation wavelengths for the two dyes.**

The FLASH-Path technique employs thin-layer dual-fluorophore staining, utilizing a nuclear-specific fluorophore (DAPI) and a lipophilic membrane tracer (DiD). As seen in Supplementary Fig. S1a, the theoretical excitation maxima for DAPI and DiD are centered at ~365 nm (UV-A) and ~640 nm (far-red), respectively. However, practical implementation in fluorescence microscopy necessitates alignment with instrument constraints. Specifically, 405 nm excitation was selected for DAPI instead of its optimal 365 nm due to the wavelength-dependent transmittance of microscope optics, particularly the objective lens. This substitution leverages the better transmittance efficiency of 405 nm light through standard fluorescence microscope components (Supplementary Fig. S1b), despite a minor Stokes shift compromise. For DiD, the native 640 nm excitation wavelength was retained to maximize fluorophore activation efficiency and minimize cross-talk with DAPI emission.

**Supplementary Note 2. Tissue surface flattening.**

The tissue surface should be flattened prior to the dye staining process to guarantee good imaging performance. The flattening procedures are described below: Prior to staining the tissue samples, the imaging surface is first flattened; the less undulation on the surface, the better the imaging results. After staining, it is necessary to adjust the imaging surface of the tissue to a position that is relatively horizontal to the focal plane of the microscope stage to reduce the imaging difficulty, as follows: first, place a layer of soft play dough on the base of the imaging component, and then stick the non-imaging surface of the tissue to the play dough; then, gently place the coverslip on the imaging surface, and press evenly with moderate pressure to ensure that the coverslip and the surface of the tissue to be examined are as tight as possible (as shown in Supplementary Fig. S2a).
